# ABCA1 activity is associated with reduced Alzheimer’s Disease risk in APOE ε4 non-carriers

**DOI:** 10.1101/2025.01.24.25321105

**Authors:** Andrés Peña-Tauber, Ricardo Hernández Arriaza, Yann Le Guen, Junyoung Park, Michael D. Greicius

**Affiliations:** Department of Neurology and Neurological Sciences, Stanford University, Stanford, CA, USA; Department of Chemistry, Stanford University, Stanford, CA, USA; Quantitative Sciences Unit, Department of Medicine, Stanford University, Stanford, CA, USA

## Abstract

**Background and Objectives:** *ABCA1* has been associated with Alzheimer’s Disease (AD) via large-scale genetic studies, but the mechanisms by which it impacts disease risk are unknown. ABCA1 catalyzes apolipoprotein lipidation in the central nervous system and is known to interact molecularly with ApoE. We explored whether variants altering ABCA1 activity depend on the presence of different *APOE* isoforms to modify AD risk.

**Methods:** We meta-analyzed European- and African-ancestry whole-genome and whole-exome sequencing datasets from the Alzheimer’s Disease Sequencing Project and UK Biobank. We used Cox hazards regression to assess the impact of rare (MAF < 1%) predicted damaging (loss-of-function, or missense with a REVEL score ≥ 0.75) variants in *ABCA1* on AD risk in *APOE* subgroups and performed *APOE* interaction analyses considering all *APOE* genotypes. We then leveraged plasma HDL levels, a phenotype known to be impacted by *ABCA1* variants, as a measure of ABCA1 activity and examined whether cumulative effects on HDL levels by *ABCA1* missense variants predict AD risk.

**Results:** Rare damaging (LOF + REVEL ≥ 75) variants on *ABCA1* increased AD risk in *APOE* ε2/ε3 and ε3/ε3 but not ε3/ε4 or ε4/ε4 cohorts. Interaction analyses indicated that damaging *ABCA1* variants increase AD risk considering all *APOE* isoforms (hazard ratio [HR] = 1.48; 95% confidence interval [CI] = 1.22, 1.79; p = 5.80E-05) and interact with both *APOE* ε2 (HR = 1.99; 95% CI = 1.30, 3.05; p = 0.002) and ε4 (HR = 0.76; 95% CI = 0.61, 0.95; p = 0.014). Predicted ABCA1 activity based on a weighted sum of HDL-associated *ABCA1* missense variants was protective against AD (HR = 0.10; 95% CI = 0.04, 0.27; p = 2.83E-06) and interacted with *APOE* ε4 to counteract this protective effect (HR = 11.66; 95% CI = 3.81, 35.66; p = 1.67E-05), while there was no significant interaction with ε2.

**Conclusions:** ABCA1 activity is protective against AD risk in *APOE* ε4 non-carriers. Distinct interactions between *ABCA1* and the main *APOE* isoforms suggest the proteins may work together to affect AD risk and motivate the development and testing of *ABCA1* therapeutics in specific *APOE* subpopulations.

## Introduction

Lipid metabolism has received attention as a potentially important pathway in the development of multiple neurodegenerative disorders.^1,2^ While many lipid-related genes including *APOE*, *ABCA7*, and *ABCA1* have now been associated with Alzheimer’s Disease (AD), the most prevalent cause of dementia, the exact role of lipid metabolism in neurodegeneration has remained elusive.^3–6^

ATP-binding cassette transporter A1 (ABCA1) is a membrane-bound lipid transporter that has been linked with AD via both common, low-effect and rare, high-effect genetic variants.^7–9^ The function of ABCA1 in peripheral lipid metabolism has been well characterized, as it is essential to produce high-density lipoprotein (HDL) particles in blood by catalyzing the efflux of phospholipids and cholesterol onto apolipoprotein A1 (ApoA1).^10^ Loss-of-function (LoF) and certain missense variants on *ABCA1* lead to decreased peripheral HDL levels, with one copy of such a variant causing familial hypoalphalipoproteinemia, and two copies producing Tangier disease, characterized by extremely low plasma HDL and an increased risk of atherosclerosis.^11^ In the central nervous system (CNS), ABCA1 appears to be important in forming “HDL-like” lipoparticles that contain apolipoprotein E (ApoE) via a process that may share molecular features of ApoA1-based lipoprotein formation.^12,13^

Lipoparticles formed by the three main isoforms of ApoE—ε2, ε3, and ε4—exhibit a stepwise decrease in lipid content mirroring their increase in risk for AD.^14,15^ This has led some to propose that the pathogenicity of each ApoE isoform in AD may be partially linked to its relative depletion in lipids and to suggest the reversal of this status as a therapeutic strategy.^16–18^ One such approach, employing a peptide that agonizes ABCA1 activity via mimicking the C-terminal end of ApoE, has been shown to increase the lipidation of ApoE particles and reverse ApoE ε4-associated pathologies in cell and animal studies.^19–22^ However, based on in vitro experiments, differences in ABCA1-driven lipoparticle formation containing each ApoE isoform remain controversial, often depending on the model system used.^19,23–26^ Further, no human data has yet characterized the relationship between *ABCA1* and *APOE* in the context of AD. Understanding this interaction at a disease level could shed light on the role of these genes, and of lipid metabolism more generally, in AD pathogenesis.

Building on the uncovered link between *ABCA1* variants and AD risk, here we examine the possibility of a genetic interaction between *ABCA1* and *APOE* by characterizing the effect of *APOE* isoforms on the risk for AD conferred by variants affecting ABCA1 activity.

## Methods

### Genetic sequencing and AD phenotyping

We analyzed individuals in the Alzheimer’s Disease Sequencing Project (ADSP) whole genome sequencing (WGS) and whole-exome sequencing (WES)^27^ and UK Biobank (UKB) WES^28^ datasets. Variants were extracted from original VCF or PLINK files and quality controlled for missingness (< 10%), Hardy-Weinberg equilibrium (p-value > 10^-8^), differential missingness between cases and controls (p-value > 10^-5^), and in ADSP, for association with sequencing platform (p-value > 10^-5^ in Fisher’s exact test) due to the inclusion of multiple cohorts. In ADSP, we included AD cases and cognitively unimpaired individuals (denoted as healthy controls), whose phenotyping included age at onset and last known healthy control age as described previously.^27^ In UKB, AD case status was set based on having a value in Date of Alzheimer’s Disease report (field 42020), and age at onset was obtained from this field minus participant year and month of birth (fields 34 and 52). All subjects without an AD report were considered healthy controls. The last known healthy age for controls was obtained from the latest censoring date in UKB (November 30, 2022) minus participant date of birth. Subjects were classified into one of five continental superpopulations—South Asian (SAS), East Asian (EAS), Amerindian (AMR), African (AFR), and European (EUR)—if they had an ancestry percentage greater than 60% as calculated by SNPWeights version 2^29^ using reference populations from the 1000 Genomes Consortium^30^. Samples were excluded if they had a genotype missingness greater than 10%.

### ABCA1 variant annotation

Variants were extracted in the *ABCA1* region (hg38 coordinates: chr9:104781002-104928246) and annotated using Ensembl Variant Effect Predictor^31^ release 107.0 with Loss-of-Function Transcript Effect Estimator (LOFTEE)^32^ and Rare Exome Variant Ensemble Learner (REVEL)^33^ plugins. Annotations were filtered to those on the canonical transcript (ENST00000374736) of *ABCA1*.

### AD association analysis

We divided each dataset (ADSP WGS, ADSP WES, and UKB WES) into ancestry cohorts and performed analyses separately in each cohort, then meta-analyzed results for high-confidence effect estimates using fixed-effects inverse-weighted variance via metafor^34^ v4.6.0 in R^35^ v4.3.3. Only cohorts with sufficient sample size for analysis (greater than 4,000 individuals) were included. We considered a result statistically significant if it had a p-value < 0.05 in the meta-analysis. We performed Cox proportional hazards regression using lifelines^36^ v0.27.8, using age at onset or last known control age as the time to event and case/control status as the right-censoring variable, to assess for association with AD. We included as covariates ten principal components produced by PC-AiR in Genesis^37^ v2.32.0, sex, and in ADSP, sequencing platform. All AD cases and healthy controls were included in ADSP. In UKB, all AD cases were included, but to reduce computational burden, controls were sampled randomly at a 10:1 ratio with cases. Related pairs of individuals up to 3^rd^ degree relatedness (kinship > 0.0442) were identified using the --make-king-table command in PLINK 2.0.^38,39^ One individual from each related pair was removed, favoring first the sample that was genotyped through WGS rather than WES, then the sample with an age at onset or last healthy control age available, and at random otherwise.

### ABCA1 rare variant burden test

We considered a set of rare variants on *ABCA1* likely to reduce gene function and which were previously associated with AD, comprised of LoF variants and missense variants with a REVEL score ≥ 0.75 (denoted LoF + REVEL ≥ 75).^9^ Variants were considered rare if they had an allele frequency (AF) below 1% in their respective cohort and all gnomAD^32^ exome and genome non-bottlenecked populations (AFR, AMR, EAS, NFE, and SAS). Variants were considered LoF if they had an IMPACT annotation of “HIGH” and were “high confidence” LoF by LOFTEE. We summed the number of alleles across selected variants for each individual (denoted “variant burden”) and performed Cox regression.

### APOE interaction analysis

To assess for differences in effect of *ABCA1* variants depending on *APOE* genotype, we calculated and compared separate effect estimates in *APOE* genotype subgroups. To robustly test for an interaction, we then examined all *APOE* genotype groups together, including four additional covariates: *APOE* ε2 dosage; *APOE* ε4 dosage; *APOE* ε2 dosage x variant burden; and *APOE* ε4 dosage x variant burden. In this analysis, we excluded individuals with an *APOE* ε2/ε4 genotype to avoid confounding by potentially conflicting effects of ε2 and ε4.

### HDL-weighted burden test

Computational tools to predict missense variant deleteriousness, such as REVEL, can be useful in genome-wide scans of gene association, but they can be noisy at an individual gene and variant level.^33,40^ We thus sought to use a more empirical method to relate estimated ABCA1 activity with AD risk, leveraging the well-described connection between *ABCA1* variants and plasma HDL. To do so, we predicted the effect of each *ABCA1* missense variant on plasma HDL values (field 30760) for participants in UKB WES and used these values to construct a score of predicted ABCA1 activity for each subject. To maximize power while still accounting for population structure and relatedness, we ran BOLT-LMM^41^ v2.4.1 on log_2_-transformed HDL values in 404,545 EUR participants in UKB with both SNP array and WES data available. SNP array data was collected as described previously.^42^ We obtained SNP array non-imputed data from all autosomes except chromosome 9 to use in the mixed model and combined them with *ABCA1* WES data for association testing, additionally covarying for age at assessment (field 21003), sex (field 31), *APOE* ε2 and ε4 dosage, use of cholesterol-lowering medication (fields 6177 for males and 6153 for females), and 40 principal components provided by UKB (field 22009). We ran BOLT-LMM with the --predBetasFile option, which includes all variants in a locus in a single model, to provide additional, more accurate estimates of single-variant beta values accounting for linkage disequilibrium. We extracted association statistics from *ABCA1* variants and kept all missense variants with an allele count of at least 10. Variants significantly affecting HDL were identified using a Bonferroni-corrected p-value < 0.05. We then used beta values from the polygenic prediction model for significant missense variants to perform a weighted burden test in AD datasets. This was done identically to the rare variant burden test described above, except the burden variable was defined as:

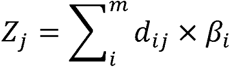

where *z_j_* is the burden variable for a subject j, *i* is each HDL-associated variant, *m* is the total number of HDL variants, *c*_ij_ is variant dosage for the given subject, and *i*_i_ is the signed beta value from HDL regression. Variants in this test were not otherwise filtered for AF. While AF filtering is necessary in the unweighted burden test to prevent the predominance of a single or few higher-frequency variants in the resulting association, in the HDL-weighted burden test, variant weight tended to be inversely correlated with AF, preventing this issue **[Supplementary Figure 2]**.

## Results

We included four cohorts in our analysis: ADSP WGS (EUR), ADSP WGS (AFR), ADSP WES (EUR), and UKB WES (EUR), for a total of 62,908 individuals passing quality control **[Table 1]**. We first examined the effects on risk of AD onset of rare LoF + REVEL ≥ 75 variants in *ABCA1* in groups defined by *APOE* genotype **[Methods]**. Through the Cox regression burden test, we replicated the finding that LoF + REVEL ≥ 75 variants on *ABCA1* increase risk for AD when considering all *APOE* genotypes together (hazard ratio [HR] = 1.30; 95% confidence interval [CI] = 1.15, 1.48; p-value [p] = 3.85E-05). Stratifying by *APOE* genotype, LoF + REVEL ≥ 75 variants increased risk in ε2/ε3 (HR = 2.40; 95% CI = 1.48, 3.91; p = 4.23E-04) and ε3/ε3 (HR = 1.62; 95% CI = 1.33, 1.97; p = 1.09E-06) groups but not in ε3/ε4 (HR = 0.99; 95% CI = 0.80, 1.21; p = 0.886) or ε4/ε4 (HR = 1.03; 95% CI = 0.67, 1.58; p = 0.882) groups. It was especially striking that *APOE* ε2/ε3 individuals saw a significant effect of *ABCA1* variants, whereas ε3/ε4 did not, despite a larger sample size in ε3/ε4 (n total = 18,978; n variant carriers = 213) than ε2/ε3 (n total = 7,334; n variant carriers = 79) **[Figure 1a]**. While we do not report effect estimates for ε2/ε2, as there were only four variant carriers in this group across all cohorts, three of these four variant carriers were diagnosed with AD, with ages at onset 57, 79, and 78, below the median age at AD onset in ε2/ε2 individuals across all cohorts (81, N = 350). Further, a log-rank test combining all cohorts indicated a significant difference in risk of AD onset in LoF + REVEL ≥ 75 *ABCA1* variant carriers compared with other ε2/ε2 subjects (p = 7.93E-09) **[Supplementary Figure 1].**

**Table 1.**
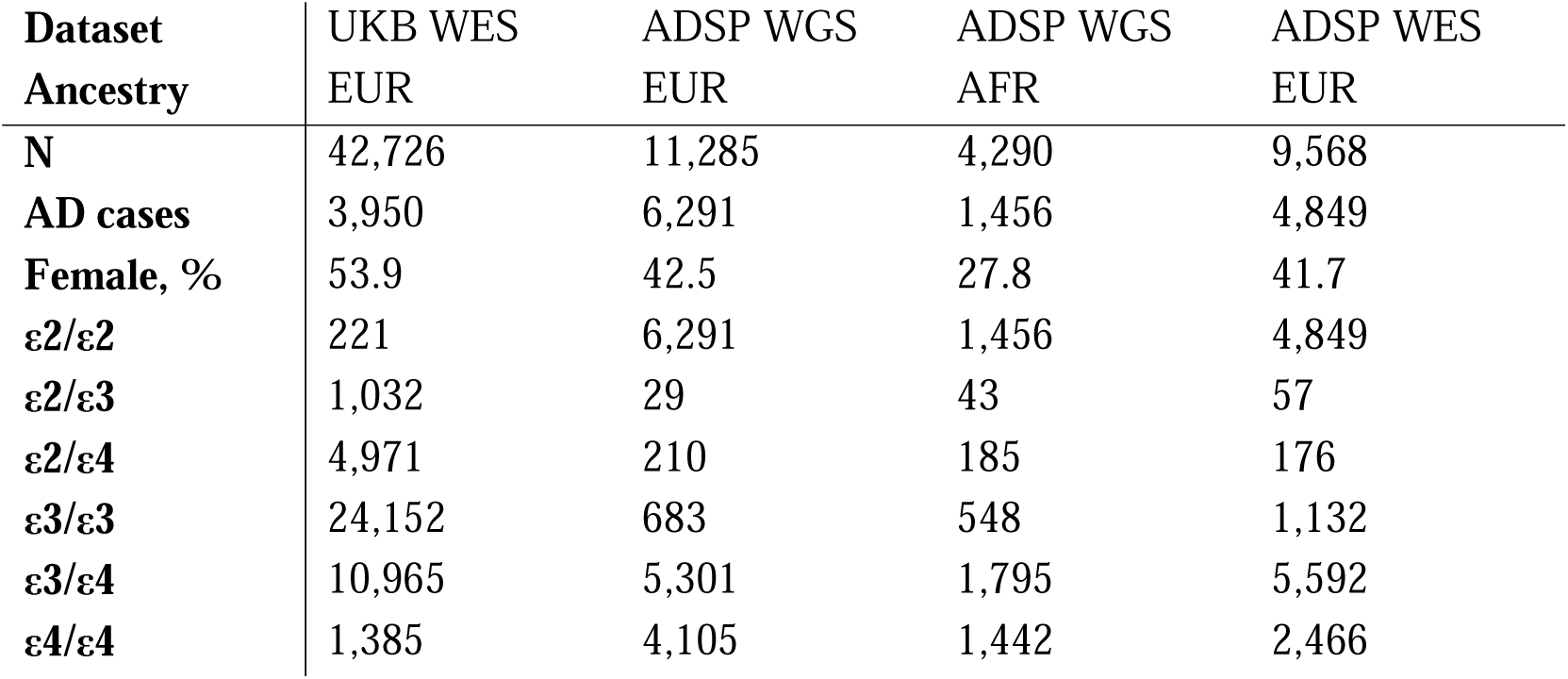
Demographic breakdown for the four cohorts analyzed in the AD burden test.

**Figure 1.**
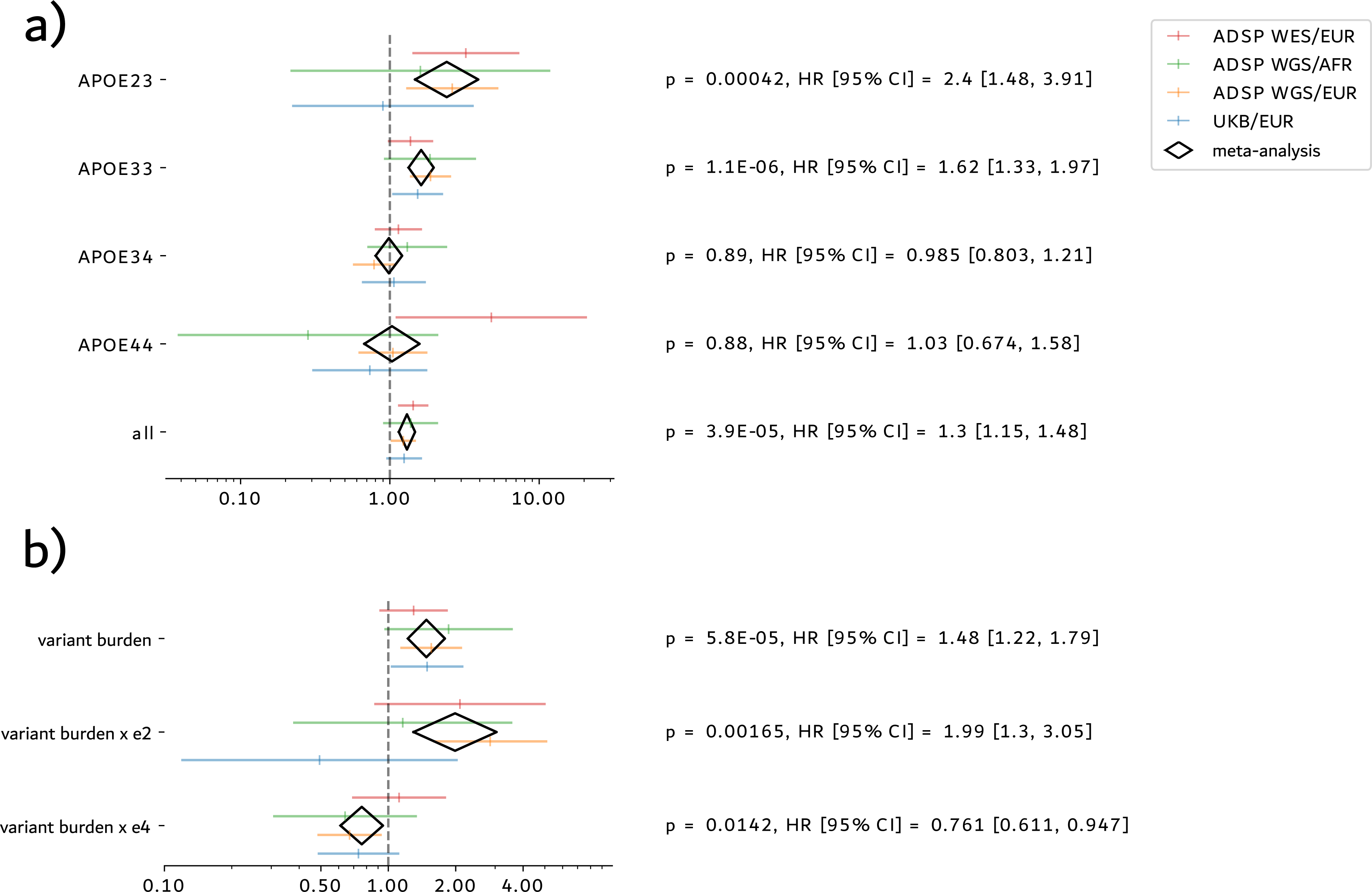
Association of burden of LoF + REVEL ≥ 75 variants in ABCA1 with AD. Bottom axes denote hazard ratios with 95% confidence intervals from Cox regression; colored lines are individual datasets, and black diamonds are the meta-analysis. (a) Results for analyses stratified by *APOE* genotype group. (b) Results for analysis including all *APOE* groups, with *ABCA1* variant dosage x *APOE* isoform dosage interaction terms.

These findings indicated that the AD risk conferred by rare, LoF + REVEL ≥ 75 variants in *ABCA1* may depend on the presence of *APOE* isoforms, with ε2 potentially increasing the magnitude of effect and ε4 reducing it. To test this hypothesis formally, we re-ran the all-*APOE* analysis in each cohort, including *ABCA1* variant burden x *APOE* isoform dosage interaction terms in the model **[Methods]**. In this analysis, we saw an AD risk effect of *ABCA1* variants by themselves (HR = 1.48; 95% CI = 1.22, 1.79; p = 5.80E-05). Further, *ABCA1* variant burden and *APOE* ε2 dosage interacted to increase AD risk (HR = 1.99; 95% CI = 1.30, 3.05; p = 0.002), and *ABCA1* variant burden and *APOE* ε4 interacted to decrease AD risk (HR = 0.76; 95% CI = 0.61, 0.95; p = 0.014), supporting the observation from the stratified analyses that both ε2 and ε4 modify the risk conferred by LoF + REVEL ≥ 75 *ABCA1* variants, in opposite directions **[Figure 1b]**.

We sought to construct a more empirically defined measure of ABCA1 activity in each subject, based on the effect of *ABCA1* missense variants on plasma HDL, to then relate ABCA1 activity to AD risk **[Methods]**. Through BOLT-LMM, we identified 35 *ABCA1* missense variants that were significantly associated with HDL levels, most of which (29) were associated with decreased HDL **[Table 2]**. We constructed a weighted sum of predicted HDL effects of HDL-associated variants in each individual, meant to capture the difference in their predicted ABCA1 activity from wild-type on a log-2 scale. Distributions of predicted ABCA1 activity were centered around zero (median = 0 – 0.014 in each cohort), and more subjects had increased (positive value, 28.7 – 84.7% in each cohort) than decreased (negative value, 0.44 – 3.1% in each cohort) predicted ABCA1 activity from wild-type. Interestingly, the African ancestry cohort had more than double the percentage of subjects with increased ABCA1 activity than any of the European cohorts **[Figure 2]**.

**Figure 2.**
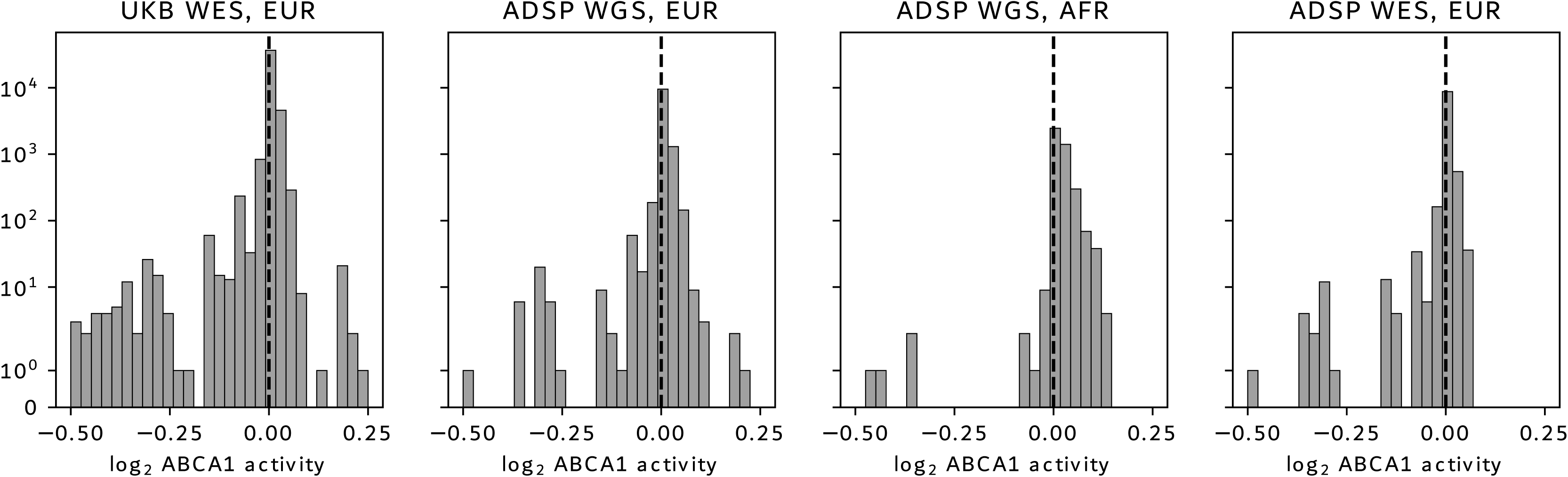
Values for weighted sums of HDL-associated ABCA1 missense variants in individuals in each cohort.

**Table 2.**
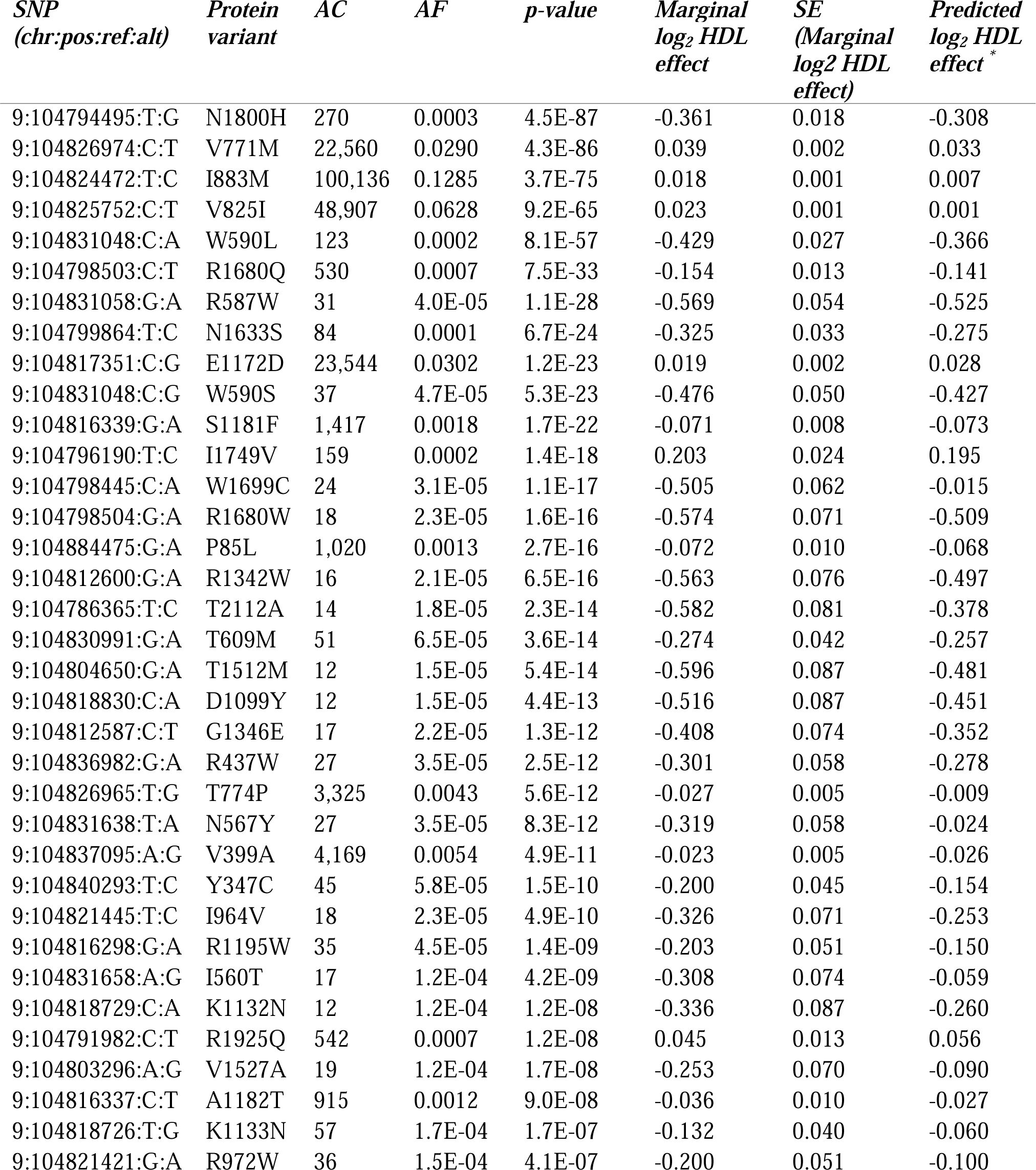
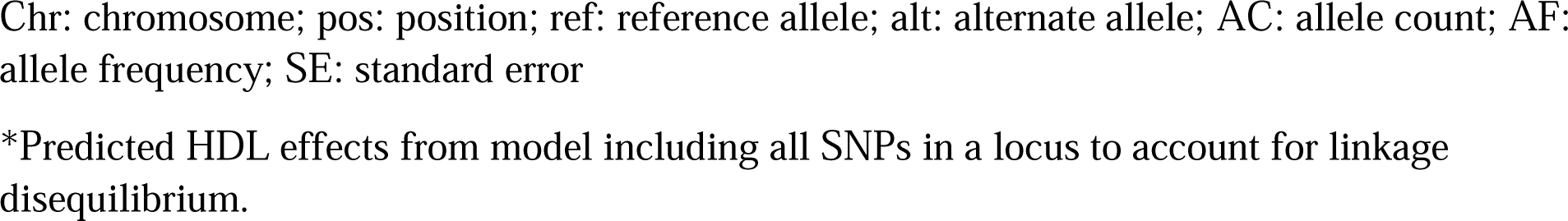
ABCA1 missense variants associated with plasma HDL levels in UKB. Variant positions are listed for human reference genome GRCh38.

| *SNP*<br>*(chr:pos:ref:alt)* | *Protein*<br>*variant* | *AC* | *AF* | *p-value* | *Marginal*<br>*log<sub>2</sub> HDL*<br>*effect* | *SE*<br>*(Marginal*<br>*log<sub>2</sub> HDL*<br>*effect)* | *Predicted*<br>*log<sub>2</sub> HDL*<br>*effect<sup>\*</sup>* |
| --- | --- | --- | --- | --- | --- | --- | --- |
| 9:104794495:T:G | N1800H | 270 | 0.0003 | 4.5E-87 | -0.361 | 0.018 | -0.308 |
| 9:104826974:C:T | V771M | 22,560 | 0.0290 | 4.3E-86 | 0.039 | 0.002 | 0.033 |
| 9:104824472:T:C | I883M | 100,136 | 0.1285 | 3.7E-75 | 0.018 | 0.001 | 0.007 |
| 9:104825752:C:T | V825I | 48,907 | 0.0628 | 9.2E-65 | 0.023 | 0.001 | 0.001 |
| 9:104831048:C:A | W590L | 123 | 0.0002 | 8.1E-57 | -0.429 | 0.027 | -0.366 |
| 9:104798503:C:T | R1680Q | 530 | 0.0007 | 7.5E-33 | -0.154 | 0.013 | -0.141 |
| 9:104831058:G:A | R587W | 31 | 4.0E-05 | 1.1E-28 | -0.569 | 0.054 | -0.525 |
| 9:104799864:T:C | N1633S | 84 | 0.0001 | 6.7E-24 | -0.325 | 0.033 | -0.275 |
| 9:104817351:C:G | E1172D | 23,544 | 0.0302 | 1.2E-23 | 0.019 | 0.002 | 0.028 |
| 9:104831048:C:G | W590S | 37 | 4.7E-05 | 5.3E-23 | -0.476 | 0.050 | -0.427 |
| 9:104816339:G:A | S1181F | 1,417 | 0.0018 | 1.7E-22 | -0.071 | 0.008 | -0.073 |
| 9:104796190:T:C | I1749V | 159 | 0.0002 | 1.4E-18 | 0.203 | 0.024 | 0.195 |
| 9:104798445:C:A | W1699C | 24 | 3.1E-05 | 1.1E-17 | -0.505 | 0.062 | -0.015 |
| 9:104798504:G:A | R1680W | 18 | 2.3E-05 | 1.6E-16 | -0.574 | 0.071 | -0.509 |
| 9:104884475:G:A | P85L | 1,020 | 0.0013 | 2.7E-16 | -0.072 | 0.010 | -0.068 |
| 9:104812600:G:A | R1342W | 16 | 2.1E-05 | 6.5E-16 | -0.563 | 0.076 | -0.497 |
| 9:104786365:T:C | T2112A | 14 | 1.8E-05 | 2.3E-14 | -0.582 | 0.081 | -0.378 |
| 9:104830991:G:A | T609M | 51 | 6.5E-05 | 3.6E-14 | -0.274 | 0.042 | -0.257 |
| 9:104804650:G:A | T1512M | 12 | 1.5E-05 | 5.4E-14 | -0.596 | 0.087 | -0.481 |
| 9:104818830:C:A | D1099Y | 12 | 1.5E-05 | 4.4E-13 | -0.516 | 0.087 | -0.451 |
| 9:104812587:C:T | G1346E | 17 | 2.2E-05 | 1.3E-12 | -0.408 | 0.074 | -0.352 |
| 9:104836982:G:A | R437W | 27 | 3.5E-05 | 2.5E-12 | -0.301 | 0.058 | -0.278 |
| 9:104826965:T:G | T774P | 3,325 | 0.0043 | 5.6E-12 | -0.027 | 0.005 | -0.009 |
| 9:104831638:T:A | N567Y | 27 | 3.5E-05 | 8.3E-12 | -0.319 | 0.058 | -0.024 |
| 9:104837095:A:G | V399A | 4,169 | 0.0054 | 4.9E-11 | -0.023 | 0.005 | -0.026 |
| 9:104840293:T:C | Y347C | 45 | 5.8E-05 | 1.5E-10 | -0.200 | 0.045 | -0.154 |
| 9:104821445:T:C | I964V | 18 | 2.3E-05 | 4.9E-10 | -0.326 | 0.071 | -0.253 |
| 9:104816298:G:A | R1195W | 35 | 4.5E-05 | 1.4E-09 | -0.203 | 0.051 | -0.150 |
| 9:104831658:A:G | I560T | 17 | 1.2E-04 | 4.2E-09 | -0.308 | 0.074 | -0.059 |
| 9:104818729:C:A | K1132N | 12 | 1.2E-04 | 1.2E-08 | -0.336 | 0.087 | -0.260 |
| 9:104791982:C:T | R1925Q | 542 | 0.0007 | 1.2E-08 | 0.045 | 0.013 | 0.056 |
| 9:104803296:A:G | V1527A | 19 | 1.2E-04 | 1.7E-08 | -0.253 | 0.070 | -0.090 |
| 9:104816337:C:T | A1182T | 915 | 0.0012 | 9.0E-08 | -0.036 | 0.010 | -0.027 |
| 9:104818726:T:G | K1133N | 57 | 1.7E-04 | 1.7E-07 | -0.132 | 0.040 | -0.060 |
| 9:104821421:G:A | R972W | 36 | 1.5E-04 | 4.1E-07 | -0.200 | 0.051 | -0.100 |

Chr: chromosome; pos: position; ref: reference allele; alt: alternate allele; AC: allele count; AF: allele frequency; SE: standard error
\*Predicted HDL effects from model including all SNPs in a locus to account for linkage disequilibrium.

We then performed Cox regression to test the association of this measure of predicted ABCA1 activity against AD risk. Here, the resulting HR reflects the proportional change in AD risk for each 2-fold change in predicted ABCA1 activity from wild-type. Predicted ABCA1 activity was associated with reduced AD risk when considering all *APOE* genotypes together (HR = 0.49; 95% CI = 0.26, 0.92; p = 0.028). Stratifying by *APOE* subgroups, the association was only detected in *APOE* ε3/ε3 individuals, in whom predicted ABCA1 activity was protective (HR = 0.06; 95% CI = 0.02, 0.15; p = 2.91E-09. Predicted ABCA1 activity was not significantly associated with AD risk in *APOE* ε2/ε3 (HR = 0.43; 95% CI = 0.03, 5.47; p = 0.515), ε3/ε4 (HR = 2.68; 95% CI = 0.97, 7.40; p = 0.058), or ε4/ε4 (HR = 1.58; 95% CI = 0.15, 16.44; p = 0.703) groups **[Figure 3a]**. These results suggested there may be *APOE* isoform differences in the effects on AD risk conferred by ABCA1 activity. Testing these differences more robustly, an *APOE* isoform dosage interaction model in the all-*APOE* group again showed that predicted ABCA1 activity was associated with reduced risk of AD (HR = 0.10; 95% CI = 0.04, 0.27; p = 2.83E-06) and that the interaction between ABCA1 activity and *APOE* ε4 dosage was associated with increased AD risk (HR = 11.66; 95% CI = 3.81, 35.66; p = 1.67E-05). We saw no significant interaction effect of ABCA1 activity with *APOE* ε2 dosage (HR = 2.12; 95% CI = 0.17, 25.82; p = 0.556) **[Figure 3b]**. Overall, these results suggested that ABCA1 activity as predicted by HDL-associated missense variants is associated with reduced AD risk in *APOE* ε2 and ε3 carriers, but that this association is weaker or nonexistent in *APOE* ε4 carriers.

**Figure 3.**
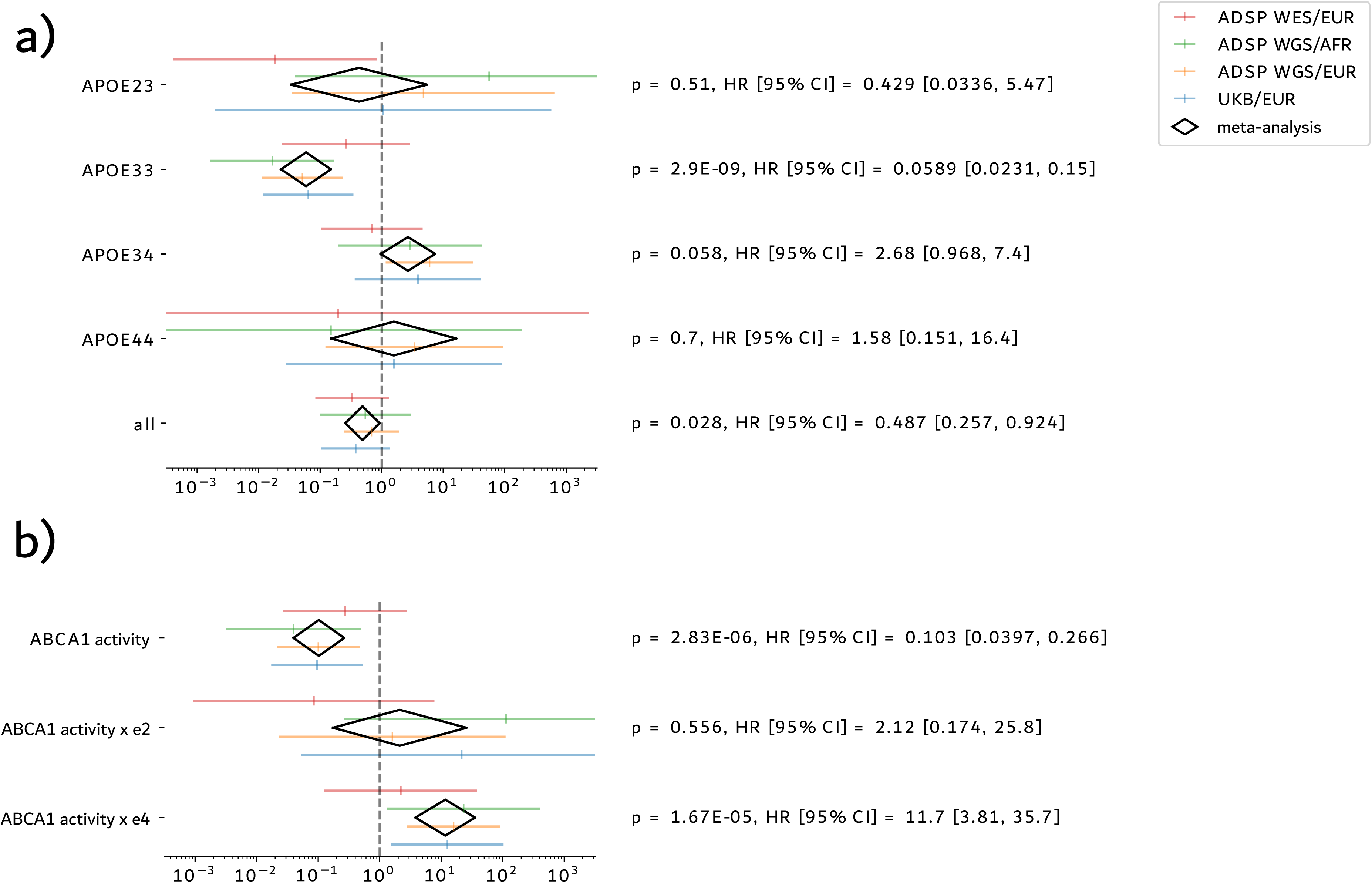
Association of predicted ABCA1 activity as reflected by a weighted sum of HDL-associated ABCA1 missense variants with AD. Bottom axes denote hazard ratios with 95% confidence intervals; colored lines are individual datasets, and black diamonds are the meta-analysis. (a) Results for analyses stratified by *APOE* genotype group. (b) Results for analysis including all *APOE* groups, with ABCA1 activity x *APOE* isoform dosage interaction terms.

We performed sensitivity analyses to validate the robustness and generalizability of the association between predicted ABCA1 activity and AD. First, we tested the impact of including only missense variants increasing (n = 6) or decreasing (n = 29) HDL levels from wild-type in the weighted sum. Weighted sums of both HDL-increasing and HDL-decreasing ABCA1 variants showed the same patterns of association as the weighted sum including all variants, suggesting the association was being driven by individuals with both increased and decreased ABCA1 activity compared to wild-type, despite there being many more individuals in each cohort with increased predicted ABCA1 activity. Second, we excluded the UKB WES dataset, as some of these individuals were used to learn HDL values, potentially introducing bias. Predicted ABCA1 activity was still associated with AD risk (HR = 0.11; 95% CI = 0.03, 0.33; p = 1.26E-04) and interacted with *APOE* ε4 dosage (HR = 11.31; 95% CI = 3.02, 42.35; p = 3.16E-04) after including only the three ADSP datasets. Third, we conducted unweighted burden tests to assess the impact of variant weights on the AD association. A burden test using simple counts of HDL-associated variants was not significantly associated with AD risk in any model, even when including only HDL-increasing or HDL-decreasing variants, supporting the weighting of variants based on their HDL effects as an important predictor of AD risk **[Supplementary Table 1]**.

In a final analysis, we explored the contribution of each variant to the association of predicted ABCA1 activity with AD in order to identify important variants for further study. For each HDL-associated missense variant, we removed the variant from the weighted sum (assigning it a weight of zero) and re-ran the association, observing changes in p-values for the weighted sum and *APOE* interaction variables. No removal of a single variant ablated the association of the weighted sum by itself or its *APOE* ε4 interaction with AD, and the removal of most variants (n = 20 out of 35) at least marginally weakened the association of the weighted sum with AD, showing that most variants included in the weighted sum are important to the association **[Figure 4a]**. We highlight five missense variants that had a large impact on the association of the weighted sum of HDL-associated variants with AD. Four missense variants led to the largest weakening of the association of predicted ABCA1 activity with AD when they were removed, by a similar amount: R587W (chr9:104831058:G:A, p = 3.00E-05 vs. 2.83E-06); N1800H (chr9:104794495:T:G, p = 2.93E-05 vs 2.83E-06); R1342W (chr9:104812600:G:A, p = 2.90E-05 vs. 2.83E-06); and E1172D (chr9:104817351:C:G, p = 2.62E-05 vs. 2.83E-06). Two of these variants can be assessed reliably by single variant statistics due to having more than 10 variant carriers across all cohorts. The variant N1800H significantly increased AD risk (HR = 2.24; 95% CI = 1.29, 3.90; p = 0.004) and interacted with *APOE* ε2 dosage (HR = 5.09; 95% CI = 1.95, 13.29; p = 8.99E-04), while it did not interact significantly with ε4 dosage but had an opposite effect direction as the interaction with ε2 (HR = 0.61; 95% CI = 0.30, 1.25; p = 0.176). This variant is the most highly associated with HDL out of all missense variants due to a large decrease in HDL levels (marginal log_2_ fold change = −0.36; 95% CI = −0.40, −0.32; p = 4.5E-87). Meanwhile, the variant E1172D was associated with decreased AD risk (HR = 0.89; 95% CI = 0.82, 0.97; p = 0.010) and interacted with *APOE* ε4 dosage (HR = 1.11; 95% CI = 1.01, 1.21; p = 0.027) but not with ε2 (HR = 1.09; 95% CI = 0.87, 1.36; p = 0.474). This variant is predicted to increase HDL levels (marginal log_2_ fold change = 0.019; 95% CI = 0.015, 0.023; p = 5.3E-23). Both N1800H and E1172D variants lead to the expected effect directions on AD risk given their effects on HDL and have significant interactions with an *APOE* isoform, thus appearing to be representative of the association we identified linking ABCA1 activity, *APOE* isoforms, and AD risk. On the other hand, the variant V771M (chr9:104826974:C:T) led to the greatest improvement in the association of predicted ABCA1 activity with AD when it was removed (p = 1.99E-08 vs. 2.83E-06). This variant is the second most highly associated with HDL out of all missense variants due to an increase in HDL levels (marginal log_2_ fold change = 0.039; 95% CI = 0.035, 0.043; p = 4.3E-86). Despite this, V771M was not associated with AD risk (HR = 1.05; 95% CI = 0.95, 1.15; p = 0.365), nor did it interact with *APOE* ε2 (HR = 1.10; 95% CI = 0.84, 1.45; p = 0.488) or ε4 (HR = 1.06; 95% CI = 0.95, 1.17; p = 0.291) dosage. Thus, this variant may act as an outlier in terms of the association between ABCA1 activity, *APOE* isoforms, and AD risk **[Figure 4b].**

**Figure 4.**
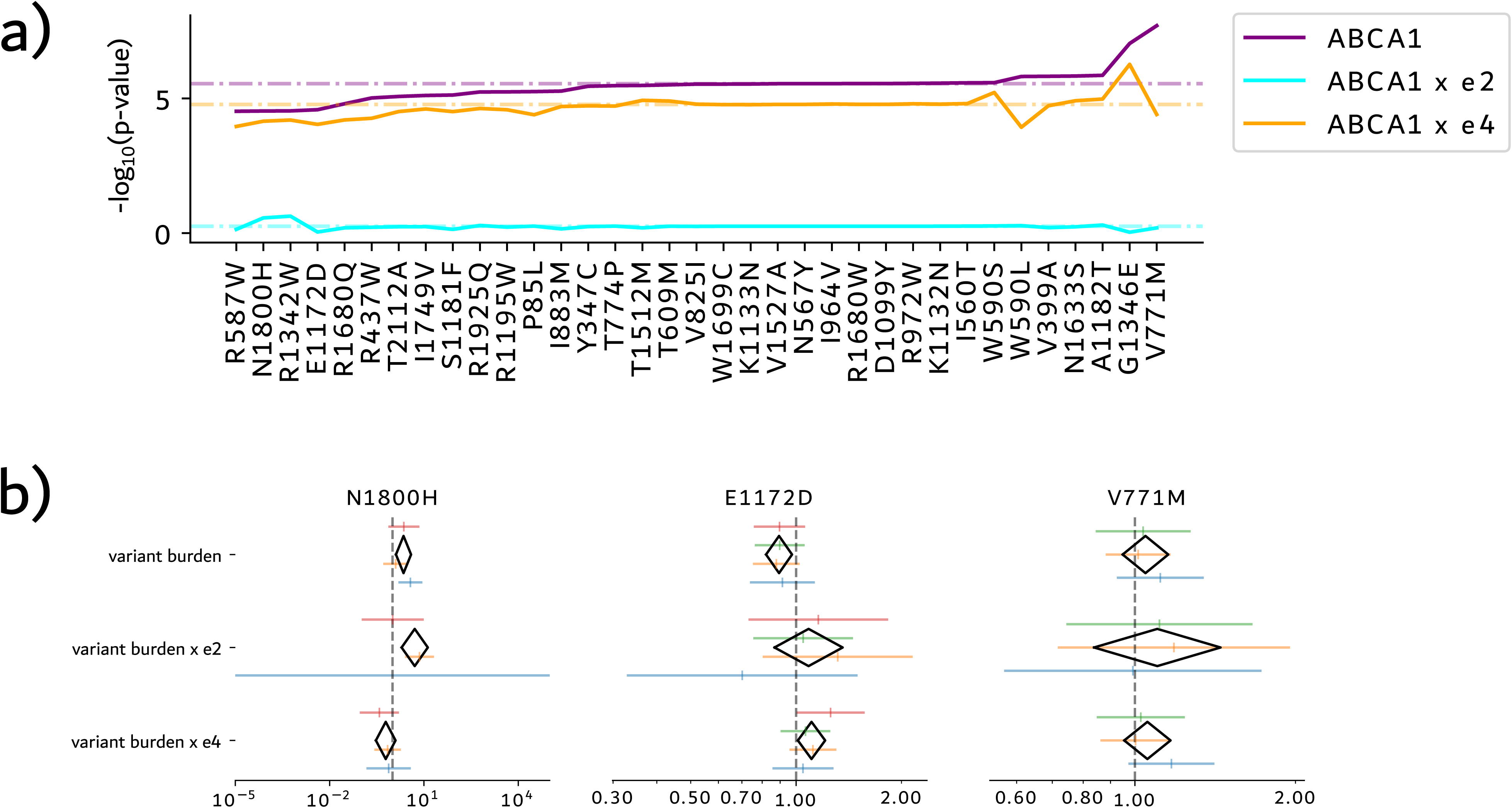
Missense variants included in the HDL-weighted burden test. (a) Contribution of each ABCA1 missense variant to the association of predicted ABCA1 activity and its interaction with *APOE* isoforms with AD. Dashed lines reflect association results for weighted sum of all variants; solid lines denote association results for weighted sum with a single variant removed. Bottom axis denotes the variant that was removed from the weighted sum. Variants are ordered according to their effects on the significance level of the weighted sum univariate term with AD. (b) AD association for three individual missense variants that had a large impact on the association between predicted ABCA1 activity and AD. Bottom axes denote hazard ratios with 95% confidence intervals from Cox regression; colored lines are individual datasets, and black diamonds are the meta-analysis.

## Discussion

The exact roles of *ABCA1* and *APOE* in AD remain unclear. While genome-wide scans such as those which identified variants in *ABCA1* as risk factors for AD are useful in nominating candidate risk genes, they do not necessarily elucidate the mechanisms by which these genes play a role in disease.^8,9^ Understanding these mechanisms requires follow-up analyses that place the risk genes within disease-relevant biological pathways and clarify the specific alterations in gene activity that lead to disease risk.

In this analysis, we identified an interaction between ABCA1 activity and *APOE* isoforms that could lend insight into the mechanisms of *ABCA1* and *APOE* in AD. First, we found a significant interaction between rare pathogenic variants in *ABCA1* (LoF + REVEL ≥ 75) and *APOE* isoforms in conferring AD risk. We found that the risk gain from these variants is strongest in ε2/ε3 individuals, who saw a 2-3 fold increase in AD risk, followed by ε3/ε3 individuals, who saw a 1-2 fold increase in AD risk, and we could not detect an effect in any group carrying an ε4 allele. An interaction model confirmed these *APOE* subgroup differences, suggesting that rare pathogenic variants in *ABCA1* are most damaging in the presence of *APOE* ε2, moderately damaging in the presence of *APOE* ε3, and least damaging in the presence of *APOE* ε4.

An additional analysis predicting ABCA1 activity based on a weighted sum of HDL effects of *ABCA1* missense variants supported the observations from rare pathogenic *ABCA1* variants. This genetic measure of predicted ABCA1 activity was protective against AD considering all *APOE* genotypes together and in *APOE* ε3/ε3 individuals, and it appeared to have no effect in ε2/ε3, ε3/ε4, or ε4/ε4 individuals. A model estimating the interaction of ABCA1 activity with *APOE* genotype suggested a difference in the AD risk effect of ABCA1 activity only in *APOE* ε4 carriers, while *APOE* ε2 carriers saw no significant difference compared to ε3/ε3. Thus, this final model indicated that *APOE* ε2 and ε3 carriers experienced an overall 4-20 fold decrease in risk for every doubling in predicted ABCA1 activity, which was largely ablated in the presence of *APOE* ε4. Importantly, while we used HDL levels to predict the effect of missense variants on ABCA1 activity, these results are not likely to mean that HDL levels themselves are causal for AD risk. Some groups have observed a correlation between HDL levels and AD or dementia risk, but Mendelian randomization analyses have been unable to support a general association between HDL-associated genetic factors and AD, making it unlikely that HDL levels themselves play a causal role.^43–47^ Instead, by including only *ABCA1* missense variants in our analysis, we refined our test to reflect the activity of a specific protein that is known to be relevant to the CNS and AD. We also ruled out the possibility of data circularity by performing a sensitivity analysis excluding UKB, which was used to estimate HDL effects. Thus, the association we identified between *ABCA1-*regulated plasma HDL levels and AD is likely an indicator that ABCA1 lipid transport activity in the CNS is related to the risk of AD, rather than plasma HDL levels themselves. However, we do not discount the possibility that some proportion of the effects we observed are related to the impacts of systemic vascular risk factors.

From both our analyses, it thus appears that ABCA1 activity is inversely correlated with AD risk—with decreased ABCA1 activity due to rare LoF variants or reduced-function missense variants increasing AD risk, and increased ABCA1 activity due to activating missense variants decreasing AD risk—exclusively, or most strongly, in *APOE* ε4 non-carriers. The genetic interactions we identified point to a potential interplay of *ABCA1* and *APOE* in AD pathogenesis. Given that alterations in ABCA1 activity seem to be impactful mainly in *APOE* ε2 and ε3 carriers without an ε4 allele, with some evidence that ε2 carriers are most affected, the protective functions of ε2 and ε3—relative to ε4—may depend on ABCA1 activity levels. These results generally support the idea that, at least partially, the AD risk effects of ApoE isoforms are related to the lipid content of the lipoparticles they form. On the other hand, AD risk in *APOE* ε4 carriers seems to be partially or wholly independent of ABCA1 activity levels. This could be explained by an intrinsic dysfunction in the ability of the ε4 isoform of ApoE to interact with ABCA1, such that increased or decreased ABCA1 activity will have a lesser or negligible effect on AD risk compared with ε4 non-carriers. This is consistent with a finding we have previously reported, in which the missense variant R251G in the lipid binding domain of ApoE protects against ε4-mediated AD risk, potentially by altering its lipid efflux capacity via interaction with ABCA1.^48^ Alternatively, the pathogenic effects of *APOE* ε4 may occur downstream of ABCA1, independently of lipid transport via this transmembrane protein. Further studies in cell and animal models will be required to understand the exact mechanisms driving this candidate pathogenic pathway. Nonetheless, our results motivate the further development of therapeutics targeting the *ABCA1* gene in AD and suggest it may be important to design *ABCA1*-targeted trials with attention to potential *APOE* subgroup effects.

While most ABCA1 missense variants in our analysis contributed to the identified link between ABCA1 activity, *APOE* isoforms, and AD risk, we call attention to three particular missense variants that had a large effect on our results and that could provide an opportunity to study the biochemical correlates of this genetic mechanism. The protein variants N1800H and E1172D led to opposite effects in terms of both apparent ABCA1 activity and AD risk. While N1800H reduced apparent ABCA1 activity and increased AD risk, E1172D increased ABCA1 activity and decreased AD risk. Further, they both interacted with *APOE* isoforms. These mutations could be studied in terms of their effects on ABCA1 localization, cholesterol efflux activity, and interactions with ApoE in diverse cell types in order to identify potential biochemical effects that lead to AD risk or protective effects. On the other hand, the protein variant V771M provides a potentially useful counterexample, given that it increases apparent ABCA1 activity and yet does not have a meaningful effect on AD risk by itself or via interacting with *APOE* isoforms. While this could be related to linkage disequilibrium confounding the HDL or AD association results, which may be confirmed by biochemical studies, it could also point to a pleiotropic protein-level effect that can be contrasted with other activating missense variants to aid in identifying the relevant pathways for ABCA1 and ApoE in AD. We also provide a catalog of HDL-associated *ABCA1* missense variants, which could be used to estimate AD risk in human carriers of each *APOE* genotype.

Collectively, our results show the potential relevance of genetic epistasis between *ABCA1* and *APOE* in driving AD risk. They underscore the importance of harnessing large genetic datasets to explore gene-gene interactions in loci that have been studied extensively, which along with structural variants and epigenetics could explain a portion of the “missing heritability” of AD and other common diseases.^49^

**Supplementary Table 1.** Sensitivity analyses of association of predicted ABCA1 activity with AD.

| Sensitivity test | ABCA1 variable | | | ABCA1 variable x $\epsilon_2$ | | | ABCA1 variable x $\epsilon_4$ | | |
| --- | --- | --- | --- | --- | --- | --- | --- | --- | --- |
|  | p-value | HR | 95% CI | p-value | HR | 95% CI | p-value | HR | 95% CI |
| HDL-increasing variants | 0.041 | 0.140 | 0.021, 0.924 | 0.236 | 22.059 | 0.132, 3694.447 | 0.013 | 11.877 | 1.674, 84.249 |
| HDL-decreasing variants | 3.45E-06 | 0.079 | 0.027, 0.231 | 0.474 | 0.344 | 0.018, 6.408 | 7.82E-04 | 10.261 | 2.637, 39.920 |
| ADSP only | 1.26E-04 | 0.106 | 0.034, 0.334 | 0.850 | 1.304 | 0.083, 20.387 | 3.16E-04 | 11.312 | 3.022, 42.349 |
| Unweighted, all HDL-associated variants | 0.144 | 0.980 | 0.954, 1.007 | 0.102 | 1.065 | 0.988, 1.147 | 0.498 | 1.010 | 0.981, 1.039 |
| Unweighted, HDL-increasing variants | 0.075 | 0.975 | 0.949, 1.003 | 0.082 | 1.069 | 0.991, 1.153 | 0.223 | 1.018 | 0.989, 1.048 |
| Unweighted, HDL-decreasing variants | 0.094 | 1.121 | 0.981, 1.282 | 0.895 | 0.974 | 0.658, 1.442 | 0.014 | 0.833 | 0.720, 0.964 |

HR: hazard ratio; CI: confidence interval

## Supplementary Figures

***Supplementary Figure 1.* Kaplan-Meier survival curves for *ABCA1* LoF + REVEL ≥ 75 variants in *APOE* subgroups, combined across all cohorts.**

***Supplementary Figure 2.* HDL effects versus allele frequencies for HDL-associated *ABCA1* missense variants.**

## Supporting information

Supplementary Acknowledgements

## Data Availability

Data will be made available after the paper is accepted by a peer-reviewed journal.

