## Supplementary figures and images for "ABCA1 activity is associated with reduced Alzheimer’s Disease risk in APOE ε4 non-carriers"

### Supplementary Figures

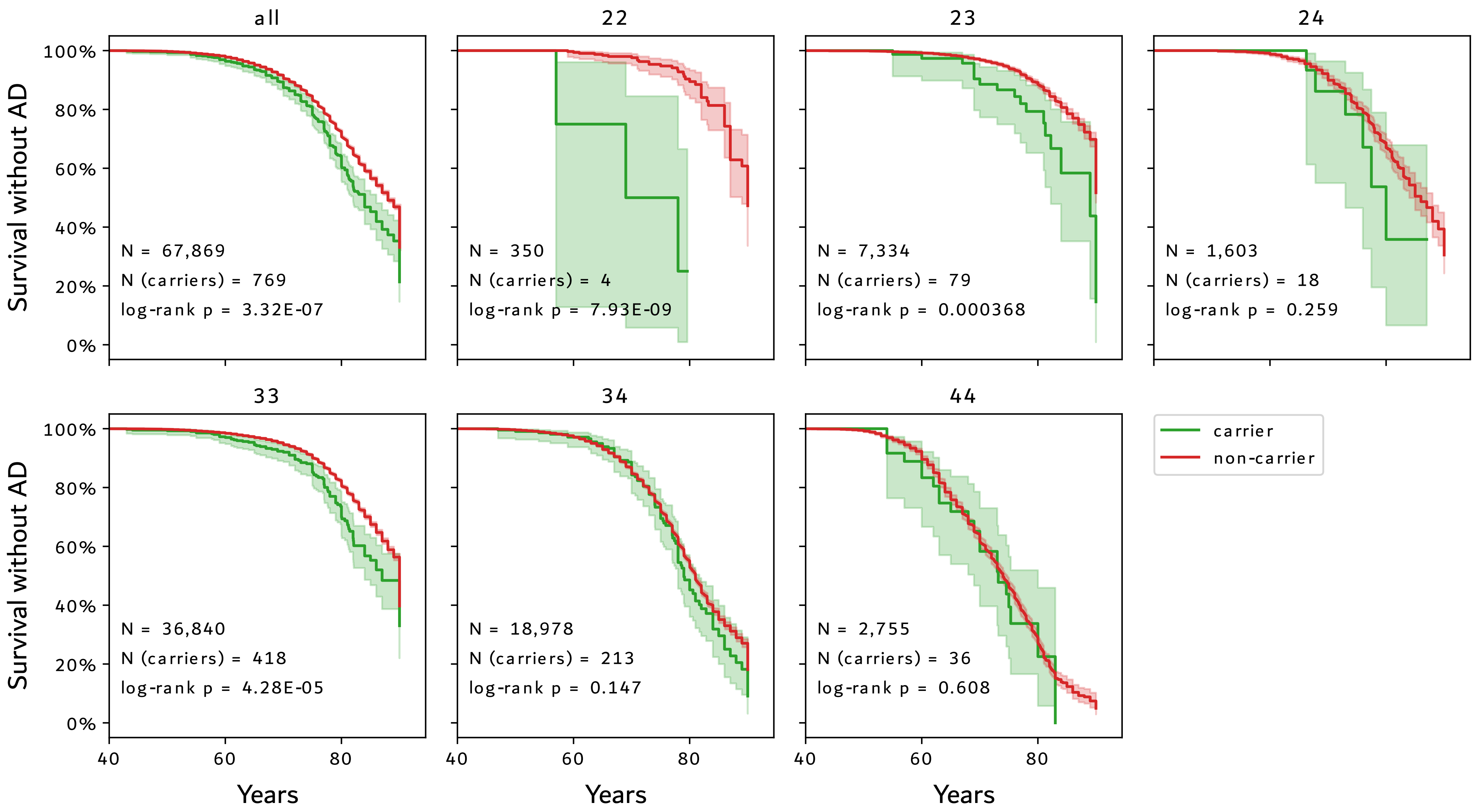

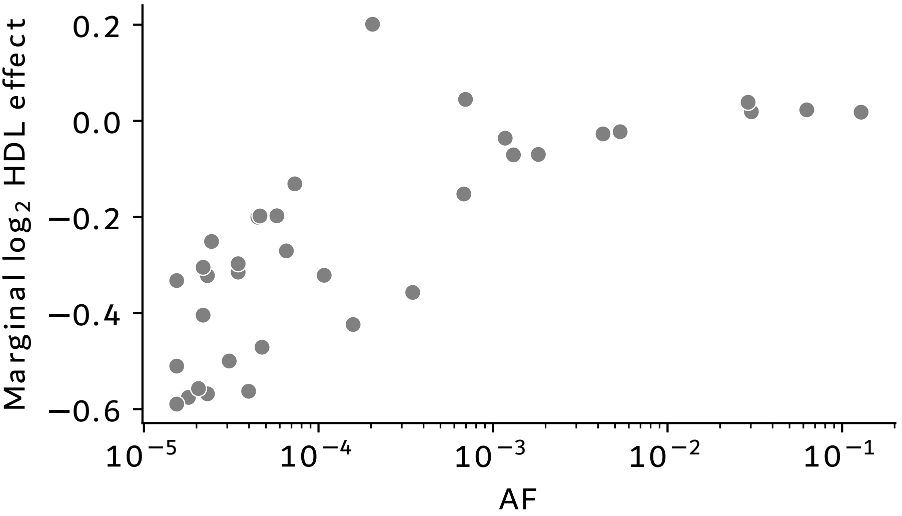
